# From Feasibility to Utility: A Meta-Analysis of Amygdala-Neurofeedback

**DOI:** 10.1101/2021.10.16.21264853

**Authors:** Noam Goldway, Itamar Jalon, Jackob N Keynan, Lydia Hellrung, Annette Horstmann, Christian Paret, Talma Hendler

**Affiliations:** Sagol Brain Institute, Wohl Institute for Advanced Imaging, Tel-Aviv Sourasky Medical Centre, Tel-Aviv, Israel; Sagol School of Neuroscience, Tel Aviv University, Tel-Aviv, Israel; Department of Psychology, New York University, New York, USA; School of Psychological Sciences, Tel Aviv University, Tel-Aviv, Israel; Department of Psychiatry and Behavioral Sciences, Stanford University School of Medicine, Stanford, CA, USA; Wu Tsai Neurosciences Institute, Stanford University, Stanford, CA, USA; Department of Neurology, Max Planck Institute for Human Cognitive and Brain Sciences, Leipzig, Germany; Laboratory for Social and Neural Systems Research, Department of Economics, University of Zurich, Zurich, Switzerland; Faculty of Medicine, University of Leipzig, Leipzig, Germany; Department of Psychology and Logopedics, Faculty of Medicine, University of Helsinki, Helsinki, Finland; Department of Psychosomatic Medicine and Psychotherapy, Central Institute of Mental Health Mannheim, Medical Faculty Mannheim/Heidelberg University, Germany; Sackler School of Medicine, Tel Aviv University, Tel-Aviv, Israel

**Keywords:** Amygdala, Neurofeedback, PTSD, Meta-Analysis, real-time fMRI, RDoC

## Abstract

Amygdala dysregulation is core to multiple psychiatric disorders. Real-time fMRI enables Amygdala self-modulation through NeuroFeedback (NF).

Despite a surge in Amygdala-NF studies, a systematic quantification of self-modulation is lacking. Amygdala-NF dissemination is further restricted by absence of unifying framework dictating design choices and insufficient understanding of neural changes underlying successful self-modulation.

The current meta-analysis of Amygdala-NF literature found that real-time feedback facilitates learned self-modulation more than placebo. Intriguingly, while we found that variability in design choices could be explained by the targeted domain, this was rarely highlighted by authors. Lastly, reanalysis of six fMRI data-sets (n=151), revealed that successful Amygdala down-modulation is coupled with deactivation of posterior insula and Default-Mode-Network major nodes, pointing to regulation related processes.

While findings point to Amygdala self-modulation as a learned skill that could modify brain functionality, further placebo-controlled trials are necessary to prove clinical efficacy. We further suggest that studies should explicitly target neuro-behavioral domain, design studies accordingly and include ‘target engagement’ measures. We exemplify this idea through a ‘process-based’ NF approach for PTSD.

## Introduction

The Amygdala, located deeply in the temporal lobe, has long been established as a main hub of emotional processing (Pessoa and Adolphs, 2010). Consistently, abnormalities in Amygdala activity and connectivity with other brain areas have been acknowledged as a transdiagnostic marker in psychiatric disorders (McTeague et al., 2020), observed for example in major depressive disorder (MDD) (Surguladze et al., 2005; Victor et al., 2010), anxiety (Brühl et al., 2014; Mochcovitch et al., 2014), borderline personality disorder (BPD) (Schulze et al., 2016) and post-traumatic stress disorder (PTSD) (Hayes et al., 2012; Mahan and Ressler, 2012). The introduction of real-time functional magnetic resonance imaging (rt-fMRI) enabled, for the first time, non-invasive self-modulation of the Amygdala through closed-looped reinforcement learning procedure termed NeuroFeedback (NF) (Sulzer et al., 2013).

In NF, real-time changes in a specific brain signal is reflected to the trainee through corresponding changes in external feedback-interface presented via auditory, visual, and/or haptic modality. The trainee is commonly instructed to change the feedback-interface in a certain way by employing mental strategies. Mental strategies associated with the desired brain signal modulation (e.g. up- or down relative to a baseline) result in a rewarding feedback, thus reinforcing neuromodulation learning. Before the introduction of fMRI, NF was employed using electro-encephalogram (i.e. EEG-NF) (Kamiya, 1969, 1968). While EEG-NF was reported to be effective in treating different neuropsychiatric disorders, the precision and validity of its effect relative to placebo remains a matter of controversy (see ref 13 for review). With rt-fMRI allowing on-line monitoring of well-localized neural signal even in deeply located areas such as the Amygdala, the interest in the application of Amygdala-NF in psychiatry has surged.

Studies so far in healthy populations have shown an effect for Amygdala-NF (using fMRI or fMRI-informed EEG) related to emotional processing including reduced difficulties in identifying and describing feelings under ongoing stress (Keynan et al., 2019), improved performance on an implicit emotion regulation task (Keynan et al., 2019, 2016), and modified emotional state (measured using Positive and Negative Affect Scale) (Liu et al., 2018). In patients, Amygdala up-modulation training using positive-memory recall was associated to improved mood and decreased depression symptoms (Young et al., 2014, 2017b, 2018), while Amygdala down-modulation training yielded alleviation of PTSD (Fruchtman et al., 2019) and BPD (Zaehringer et al., 2019) symptoms. With that, some studies suggest less straight-forward conclusions. For example, several investigations demonstrated mixed results with respect to Amygdala modulation abilities, either within the experimental group (Marxen et al., 2016; Paret et al., 2018), or when compared against active controls (46). This is true also in the case of clinical outcomes. For example, in chronic pain, the control group demonstrated similar clinical effect to the Amygdala-NF at the end of the NF training protocol (Goldway et al., 2019). Although it seems that the majority of Amygdala-NF studies point to an effect, a systematic quantitative summary of existing evidence is still needed. Such a summary could help sort out sources of learning variance and difference in effect sizes, and through that inform about expected robustness of the neuromodulation effect in association to study parameters. Additionally, several studies have examined neural modifications following Amygdala NF procedure. Findings point to simultaneous enhancement in functional connectivity between the Amygdala and medial prefrontal cortex (mPFC), premotor cortex and rostral anterior cingulate cortex (Keynan et al., 2019; Paret et al., 2016c; Zotev et al., 2013) as well as post training decrease in Amygdala reactivity to emotional stimuli in patients (Misaki et al., 2018a; Nicholson et al., 2017; Paret et al., 2016a; Young et al., 2018; Yuan et al., 2014; Zotev et al., 2018). However, in most cases, it is yet unclear what aspect of these modifications could be attributed to individual success in neuromodulation. It is well recognized that not all participants are able to regulate their neural signals to a similar extent, hence unveiling the mechanism of such individual difference could further improve NF utilization (Alkoby et al., 2017; Anna Weber et al., 2020; Kadosh and Staunton, 2019).

The current study summarized available published data of Amygdala-NF, while addressing three issues related to Amygdala-NF feasibility and/or utilization:

1. **Effect size of Amygdala self-modulation:** Two types of evidence are needed to support the premise of volitional control over brain signals. (a) NF training leads to learned self-modulation of the targeted brain signal and (b) Such learning results in neurobehavioral or clinical change. To examine these effects we conducted a meta-analysis of existing Amygdala-NF studies in healthy and clinical populations, while considering reported effects of neuromodulation and clinical outcome.
2. **Design parameters and Amygdala related processes**: Several recent reviews highlighted the need for an organizing framework to guide design choices (Paret et al., 2019; Thibault et al., 2018). Here, we tested whether the Research Domain Criteria (RDoC) framework (Cuthbert, 2014) could be used to explain experimental design choices in Amygdala-NF studies. For this, we summarize design parameters used in research so far and examined them it in light of positive and negative valence processing; major processing domains of the Amygdala (Beyeler et al., 2018; O’Neill et al., 2018).
3. **Neural mechanism of successful Amygdala self-modulation:** Studies examining the neural mechanism of Amygdala-NF so far (Keynan et al., 2019; Paret et al., 2016c, 2016a; Zotev et al., 2013), mostly analyzed entire cohorts without considering individual success in neuromodulation. This precluded differentiation between general effort to extract modulation and learning processes. Using a cross-lab large fMRI-NF data set while accounting for Amygdala modulation success (defined in a similar manner across studies), enabled us to unveil the neural mechanism that underlie effective volitional neuromodulation of the Amygdala.

## Methods

### 1. Effect size of Amygdala self-modulation

First, the data-set was defined by searching exiting studies of Amygdala-NF guided by BOLD activity or its EEG signature (1a. below). Second, the effect sizes were quantified by performing meta-analyses assessing neuromodulation, learning and clinical outcome (1b. below).

#### a. Data search

The search was conducted according to the Preferred Reporting Items for Systematic Reviews and Meta Analyses (PRISMA) guidelines (Liberati et al., 2009). Available published Amygdala-NF studies were identified until January 16th, 2021 through a search within two databases: EMBASE and MEDLINE, using the terms “Amygdala” AND “neurofeedback”. In addition, relevant reviews were used to identify articles that might have been missed in the database queries. Three trained investigators independently reviewed titles and abstracts. Studies were excluded as not being relevant in a consensus meeting (N.G, J.N.K, and T.H). The following criteria for inclusion were implemented: 1. An original research article, 2. A minimal number of five participants, 3. NF intervention studies are based on either Amygdala-fMRI (with no additional brain targets) or its EEG signature. For comparability, we excluded studies that used functional connectivity matrices, MVPA NF or feedback that is based on multiple NF probes (for example, two ROIs simultaneals). We did include studies that involve an EEG probe that is based on Amygdala-BOLD activation known as the Amygdala Electrical FingerPrint (Amyg-EFP) (Meir-Hasson et al., 2016, 2014). The Amyg-EFP was found to correlate with simultaneously acquired right Amygdala BOLD activity in a separate group than the one used for model development (Keynan et al., 2016). More so, Amyg-EFP-NF training (relative to sham control) resulted in better Amygdala self-modulation as measured by Amygdala-fMRI-NF (Fruchtman et al., 2019; Keynan et al., 2019, 2016). In light of these validation results, studies using the Amyg-EFP probe were included in the meta-analysis. If multiple publications from the same data were available, we included only the one with the most detailed information regarding the NF procedure. Altogether, 33 publications originating from 24 studies met the inclusion criteria (see supplementary Figure 1) with a total of 535 participants in the experimental condition and 251 in control conditions. The summary of these studies can be found in Table 1. Graphical discerption of the procedural aspects of the included studies (population type, sample size, number of sessions) can be found in Figure 2.

**Table 1.**
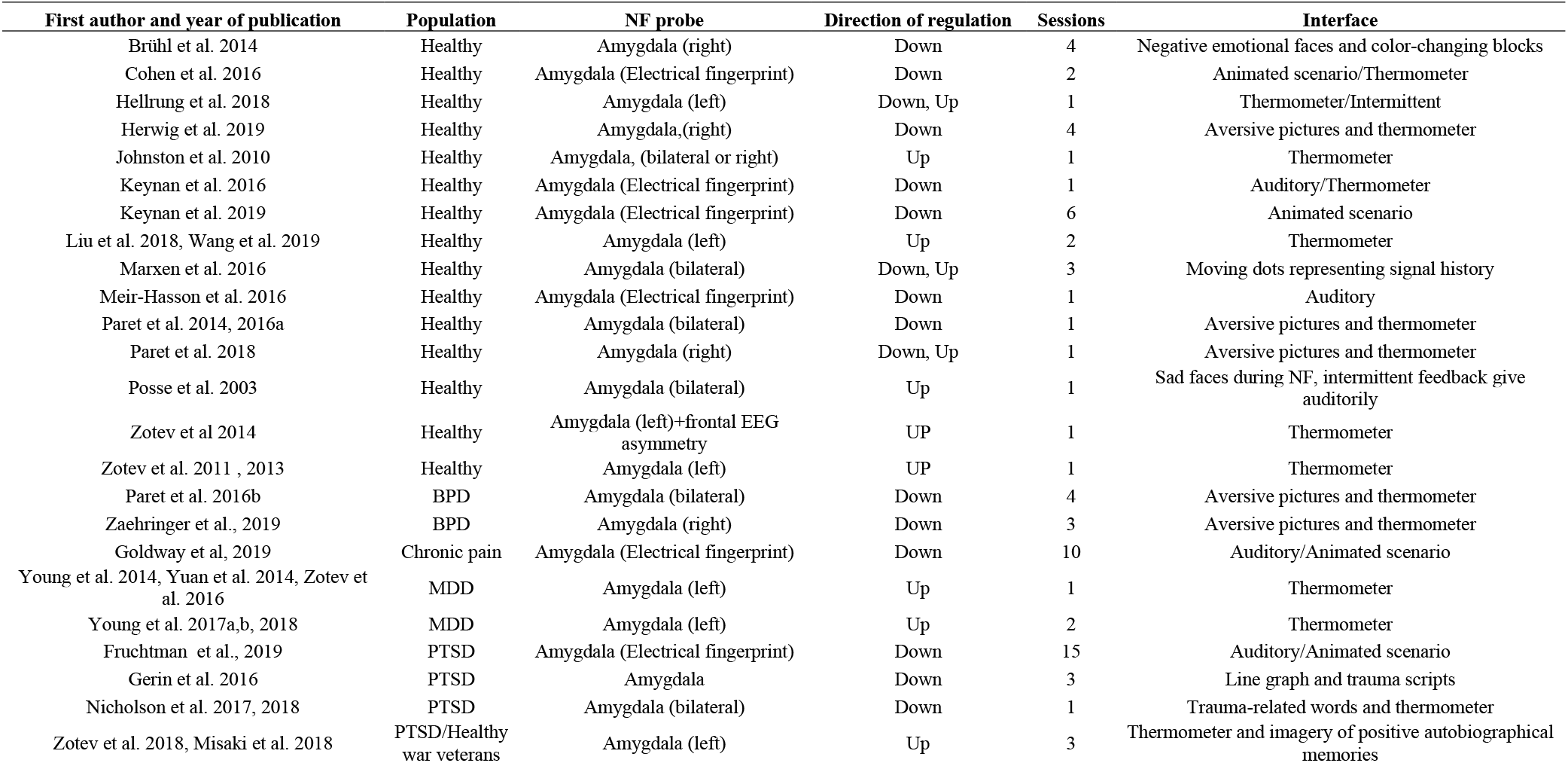
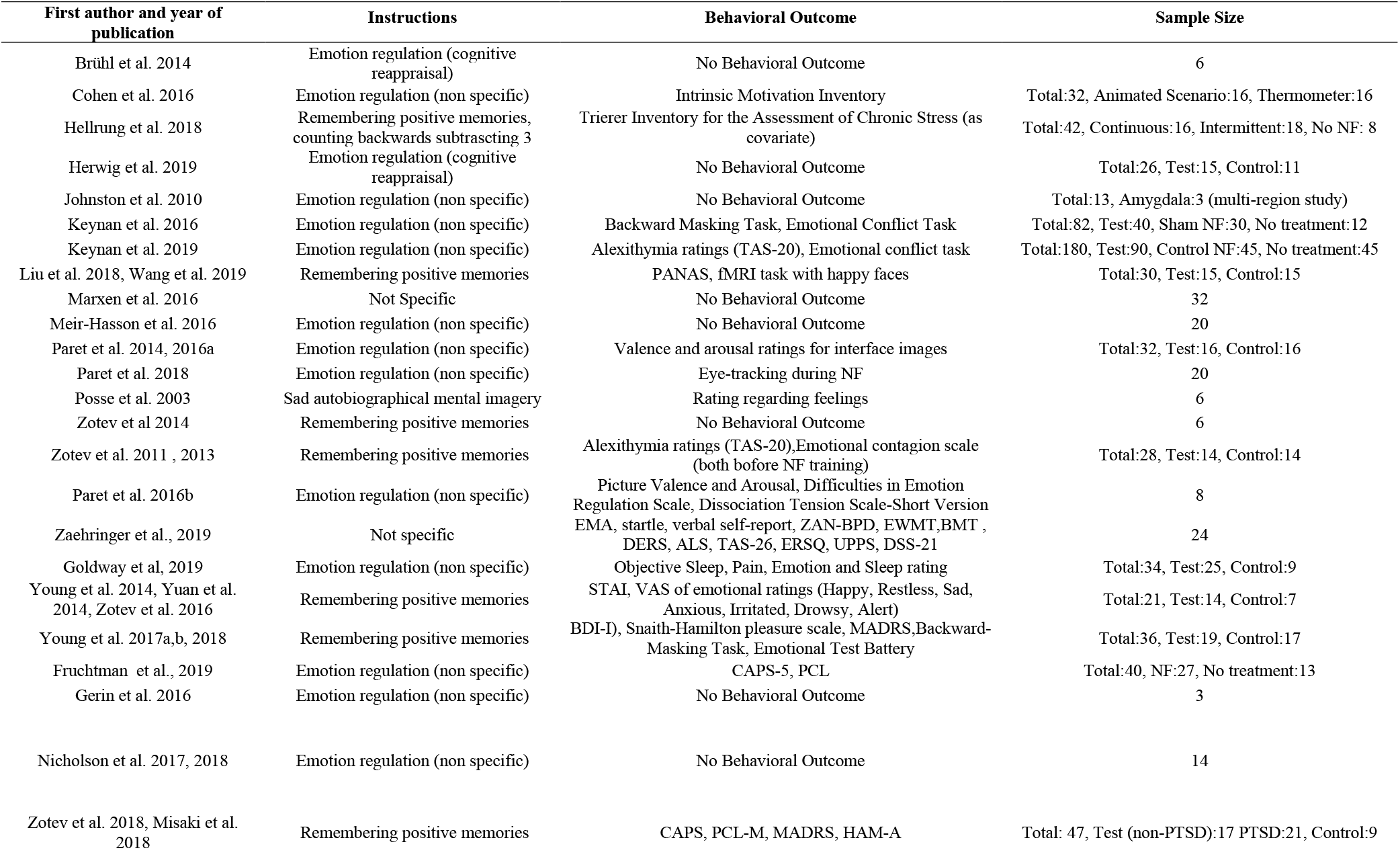
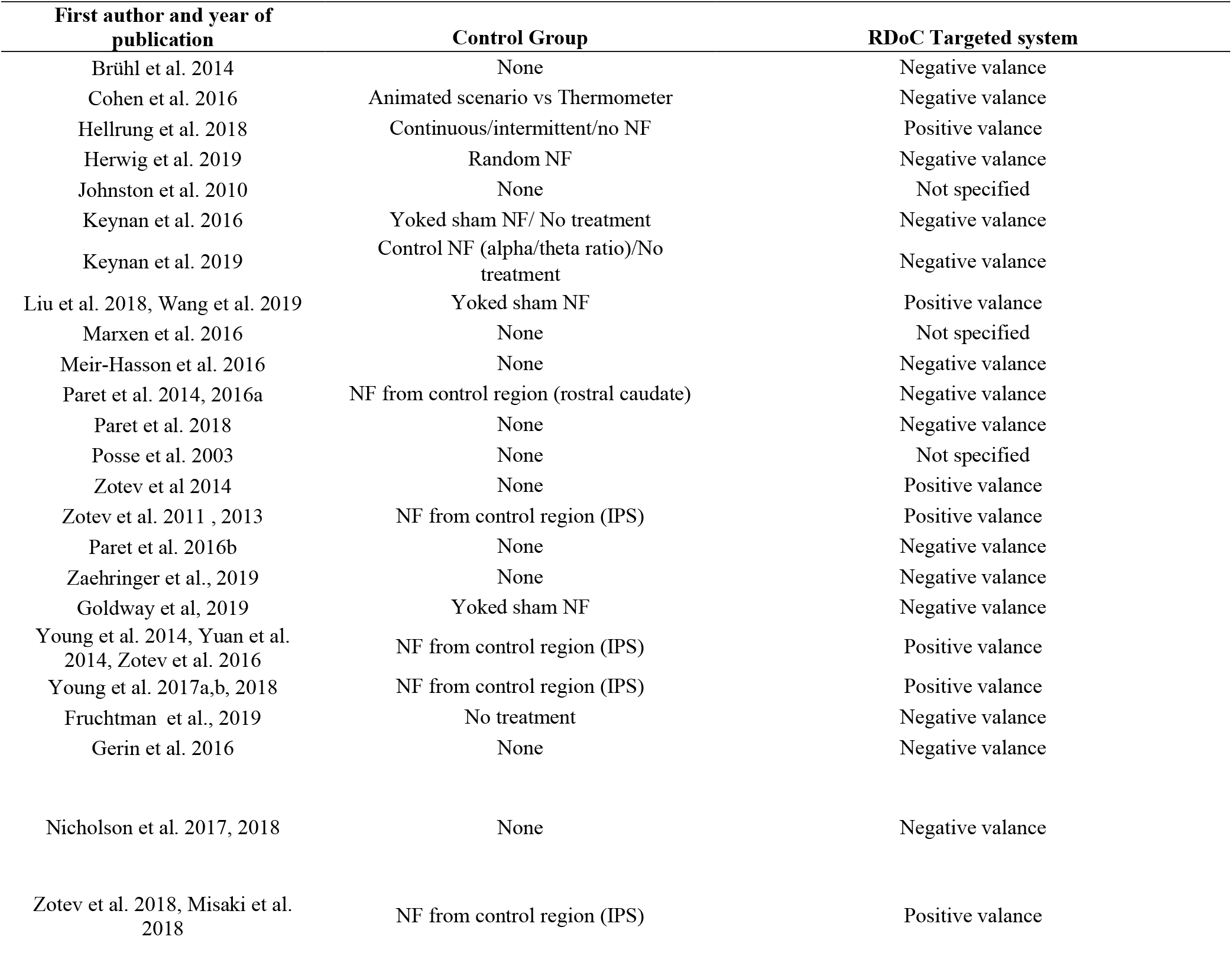

#### b. Quantitative assessment of Amygdala self-modulation

Papers were included in this part of the analysis only if they explicitly reported T or F values and degrees of freedom or provided source data electronically. In case these were not reported or extractable from the reported data in the paper, we reached out to the corresponding authors, requesting this information. If these data were supplied, the paper was included in the final analysis (see supplementary Figure 1). For all sources that met inclusion criteria, effect sizes, 95% confidence intervals, and their standard errors were calculated using “dmetar” software package (https://bookdown.org/MathiasHarrer/Doing_Meta_Analysis_in_R/). Based on current recommendations (Cuijpers, 2016), all indices were evaluated by pooled random effects (Borenstein et al., 2011). For standardization, all effect sizes were transformed to Cohen’s d metric (Cohen, 1988) using conventional formulas (http://core.ecu.edu/psyc/wuenschk/SPSS.htm).

Three types of contrasts were evaluated for effect sizes either for within or between group effects (total of six): 1. *<u>Neuromodulation</u>* <u>effect</u> was defined as the difference in Amygdala activity between *baseline* and *regulate* conditions across all experimental sessions/runs. The *regulate* condition refers to blocks where participants actively attempted to regulate the Amygdala signal. The *baseline* condition refers to blocks where participants were instructed not to perform modulation. This effect size was pooled from all included studies and refers to the Amygdala-NF experimental group only. The between group effect size was based on the difference in neuromodulation between Test and Control groups. This analysis was performed only for studies including an active control condition (i.e. NF from a control region or from shame signal). 2. *<u>Learning</u>* <u>effect</u> was defined as the difference in *Neuromodulation* between the first and last NF sessions. This was only possible to calculate for studies that included more than one NF session. The between group effect size was based on the difference in learning between Test and Control groups. This was only possible to calculate for studies that included more than one session and active control condition. 3. *<u>Clinical outcome</u>* <u>effect</u> was defined as the difference in the main clinical outcome post vs pre NF training, this index was pooled only for studies involving clinical population. The between group effect size was based on the differences in *clinical outcome* between the Test and Control groups. Control groups were either active or no-treatment. The summary statistics of the meta-analysis included two indices. First, overall effect; a weighted mean pooled from all studies included in the analysis. The coefficient wight for each study was calculated based on the confidence interval of the estimated effect which is influenced by the study’s sample size. Secondly, the I^2^ index was calculated, accounting for the variability in the effect sizes which is not caused by sampling error (Higgins and Thompson, 2002).

### 2. Design parameters and Amygdala related processes

From each study in Table 1, we extracted the following parameters: direction of neuromodulation (up/down, both), regulate instructions (non-specific emotion regulation, cognitive reappraisal, positive memory recall, sad autobiographic memories, no-instructions), clinical/behavioral outcome measures (self-report rating, clinical evaluation, behavioral task, no-outcome), neuromodulation probe (right, left, bilateral Amygdala BOLD or EFP), control condition (sham, control region, none) as well as the relatedness to positive or negative valence systems that was targeted in the study. The last parameter was extracted based on the theoretical rational outlined in the introduction of the studies. In cases where the positive, nor negative valance systems were referred to in the study rational we did not refer to this parameter in our summary.

Extracted data was further used to evaluate whether the RDoC framework that could explain experimental choices. Specifically, we examined if this framework could explain the between-study distribution of three theoretically relevant design choices (i.e. direction of modulation, NF probe, and instructions), better than the clinical diagnosis of the study population. To this end, we clustered design choices based on the RDoC neurobehavioral domain (positive or negative valence system) that was targeted in each study and compared it, as a control analysis, to clustering based on clinical diagnosis of the study population (Healthy, BPD, Chronic pain, MDD or PTSD). Distribution was then tested for being different than chance (chi-square test) per each design parameter.

### 3. Neural mechanism of successful Amygdala self-modulation

To address this objective, we obtained data from six Amygdala-fMRI-NF studies (some of this data was previously analyzed. (See, Hellrung et al., 2018; Keynan et al., 2019, 2016; Paret et al., 2014), collected by three different labs: Central Institute of Mental Health in Mannheim (n=16), Max Planck Institute for Human Cognitive and Brain Sciences (n=33), and Sagol Brain Institute, Tel Aviv Medical Center (n=102). All studies included healthy individuals that participated in a single session of Amygdala-fMRI-NF for down-modulation with a visual feedback-interface (for specific acquisition methods see supplement material). Raw NIFTI or DICOM images were subjected to a uniform processing pipeline using SPM12 (Penny et al., 2011) and MATLAB 2018a (MathWorks, Inc), including motion correction to mean functional image, co-registration to anatomical image, normalization to MNI space, and spatial smoothing with a 6mm full width half maximum gaussian kernel (for more details per data set see original publications and supplement). In addition to task regressors for *regulate* and *baseline* conditions and six motion regressors were included in the GLM. First level contrast maps of the *regulate* vs *baseline* contrast were used to perform second-level analysis (random effects group-level analysis) of the *regulate* vs *baseline* contrast. To control for variance resulting from multi-center acquisition, three “center” nuisance regressors were included in the GLM analysis, corresponding to the three different acquisition sites.

To examine different activation patterns in participants who performed better or worse in down-modulating their Amygdala, the dataset was divided into two sub-groups (termed hereby Successful and Unsuccessful, respectively). The division was performed by extracting mean beta values in the targeted Amygdala region (left, right or bilateral, according to the probe used in the original study) for each participant, and splitting the dataset according to beta<0 (Successful), or beta>0 (Unsuccessful) for all participants of all included studies. This resulted in a Successful modulators group (n=72, mean Amygdala beta=-0.51, SD=0.62), and an Unsuccessful modulators group (n=79, mean Amygdala beta=0.44, SD=0.4). Second level analyses of the *regulate*>*baseline* contrast were performed for: The whole sample (n=151), Successful modulators (n=72) and Unsuccessful modulators (n=79). To further characterize success related neuromodulation we performed an additional second level analysis of the Successful modulators group, including a continuous “success” covariate composed of the mean Amygdala beta value for down *regulate* > *baseline* derived from for the *regulate* vs *baseline* contrast, meant to identify regions whose activation is modulated along with the Amygdala during successful down-modulation.

## Results

### 1. Effect size of Amygdala self-modulation

The meta-analysis indicated a large effect size for *Neuromodulation* measured by within-experimental-group differences in Amygdala activation during *regulate* relative to *baseline* across 20 studies (SMD=0.87, 95%-CI=0.59-1.1, T=6.46, p<0.0001) (Figure 1a). The *Neuromodulation vs Placebo* effect-size was smaller than the within-group effect, yet nonetheless significant (SMD =0.56, 95%-CI=0.23-0.90, T=3.84, p<0.005) (Figure 1b). *Learning* effect size measuring within experimental-group differences in *Neuromodulation* learning effect (last vs first NF session) revealed medium effect size (SMD=0.55, 95%-CI=0.24-0.86, T=3.91, p<0.005) (Figure 1c). Between group assessment (Test vs Control) revealed a medium-strong effect size (D=0.69, 95%-CI=0.39-0.99, T=5.29, p<0.001) (Figure 1d).

**Figure 1:**
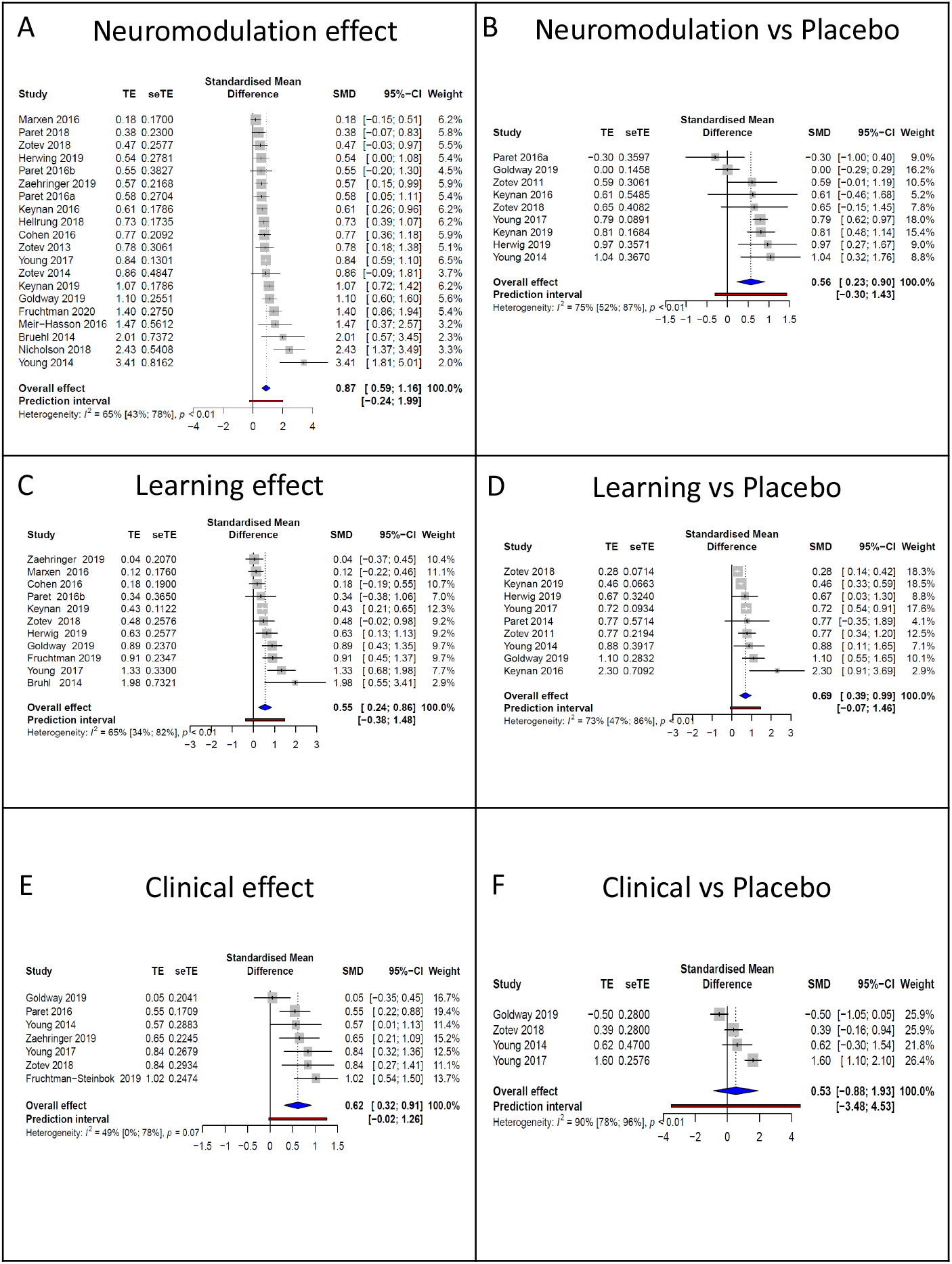
Effect size of Amygdala neuromodulation and clinical effects across the literature. (a) Forest plot illustrating the *neuromodulation* effect size in the Amygdala for *regulate* vs *baseline* conditions within the experimental group. (b) Forest plot of the contrast *regulate* vs *baseline* between experimental and control groups. (c) Forest plot of the difference in neuromodulation between the first and last NF sessions within the experimental group. (d) Forest plot of the difference in *Learning* between the experimental and control groups. (e). Forest plot of the difference in the main clinical outcome post vs pre neurofeedback within the experimental group. (f) Forest plot of the differences in Clinical effect between the experimental and control groups. The ‘Weight’ column indicates the contribution of the paper. The overall effect appears as a blue diamond-shaped object at the final row. The prediction interval indicates the range in which further observations are likely to occur. TE: estimated effect (of modulation), seTE: Standard error of estimate, SMD: summary measure for effect size Cohen’s d, CI: confidence interval.

Heterogeneity factor *I*^2^ indicated significant between-study variance for all the above mentioned indices (*Neuromodulation* within, *I*^2^= 65%, 95%-CI=43%-78%, p<0.01; *Neuromodulation vs placebo, I*^2^= 75%, 95%-CI=52%-87%, p<0.01; *Learning within, I*^2^= 65%, 95%-CI=34%-82%, p<0.0; *Learning vs placebo, I*^2^= 73%, 95%-CI=47%-86%, p<0.01). To follow up on the sources of this variability, we conducted a subgroup analysis (Borenstein et al., 2011), examining the contribution of study design to the between-study variance in effect size (see supplementary material). Importantly, this analysis revealed no significant results (all p>0.15), indicating that this variability could not be attributed to one specific design factor.

The current literature includes a relatively small number of studies reporting clinical outcomes (7 total studies, 4 of which were placebo-controlled). Following recent guidelines (Ioannidis et al., 2008) we nonetheless conducted a meta-analysis of the reported effects. *Clinical effect* was defined as the difference in main clinical outcome post- vs pre-treatment within the experimental group. For studies including a control group, *Clinical vs Placebo* effect size was further extracted testing the difference in *Clinical effect* between the experimental group and control. The main outcomes were: Fibromyalgia; Goldway et al (Goldway et al., 2019) - pain compound score, MDD; Young et al a (Young et al., 2014) - Profile of Mood States (McNair, 1971), Young et al b (Young et al., 2017c) - Montgomery-Asberg Depression Rating Scale (Montgomery and Åsberg, 1979), BPD; Paret et al (Paret et al., 2016b) - Difficulties in Emotion Regulation Scale (Gratz and Roemer, 2004), Zaehringer, et al (Zaehringer et al., 2019) - Zanarini rating scale for BPD (Zanarini, 2003), PTSD; Zotev et al (Zotev et al., 2018) and Fruchtman et al (Fruchtman et al., 2019) - the Clinician-Administered PTSD Scale (Weathers et al., 2013). The analysis revealed a medium effect size for the *clinical effect* within the experimental group (D=0.62, 95%- CI=0.32-0.92) with a marginally significate between-study heterogeneity factor *(I*^2^= 49%, 95%-CI=0%-78%, p=0.07) (Figure 1e). Similarly, *Clinical vs placebo* effect could also be described as medium (D=0.53, 95%-CI=-0.88-1.93) however, with substantial heterogeneity (*I*^2^= 90%, 95%-CI=78%-96%, p<0.01) (Figure 1f).

### 2. Design parameters and Amygdala related processes

The distribution of design choices in each of the examined category is illustrated by a colored box per study in Figure 2b. The figure points to a vast variability concerning basic design choices in the current literature. For Training Instruction the most common practice was emotion modulation and retrieval of positive memories, used in about a quarter of the studies. Only two studies provided no instructions. For Outcome Measures about a third of the studies didn’t assess outcome, and the rest are equally distributed for behavioral tasks and self-report or clinical scales. For NF Probe half of the studies targeted the right Amygdala (half of them via BOLD and half via EFP). The rest applied left or bilateral. For Control Conditions, about a third did not include a control condition. Control conditions were either a different region or yoked shame. For Direction of Modulation, down- regulation was the most common practice, yet a third applied up-modulation and only a few conducted both.

**Figure 2:**
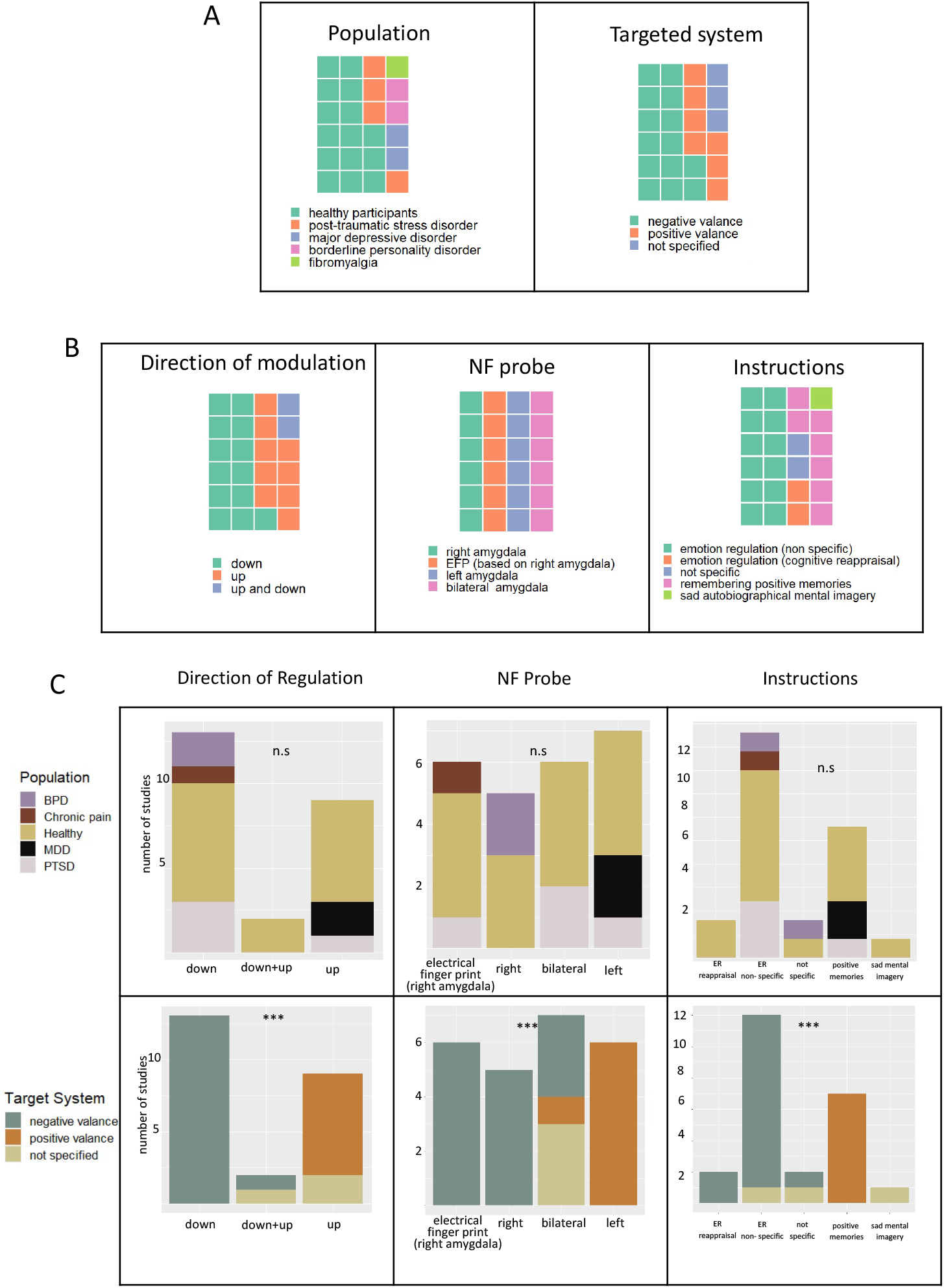
Distribution of Amygdala-NF design parameters. (a) the distribution of study population and of the neurobehavioral target system. Left panel: The majority of studies were done on healthy participants (n=363), 4 studies involved PTSD (n=65), 2 studies involved MDD (n=33), 2 studies involve BPD (n=25), and one study of chronic pain (n=25). Right panel: The negative valence system is the most commonly targeted cognitive system, while positive valence is targeted at just over a quarter of studies. (b) distribution of design parameters expected to be derived by theoretical considerations. Left panel: Most commonly, participants are instructed to down-modulate their Amygdala, but a significant proportion of studies aimed for up-modulation. Only in two methodological studies, participants were instructed to both up- and down-regulate their Amygdala activity. Middle panel: Dominance is observed for targeting the right Amygdala, with half of the studies targeting either this probe directly or using Amyg-EFP. Right panel: Most dominantly, emotion regulation related instructions were provided to facilitate Amygdala-NF, while retrieval of positive memories was used in about a quarter of the studies. Additionally, induction of sad memories were used. Two studies provided no instructions. (c) Explaining the variability in Amygdala-NF design parameters. Top row: clustering studies based on the clinical diagnosis that was targeted in the NF experiment. Bottom row: clustering studies based on the neurobiological processes that were targeted in the NF experiment. Left column: the direction of modulation reflected the desired change in Amygdala modulation compared to baseline. Middle column: Amygdala-NF probe that was targeted in the NF experiment. right column: type of instructions that were provided to the trainees to achieve Amygdala modulation. p-values represent the result of the chi-square test.

As seen in Figure 2c, grouping studies by the targeted RDoC neurobehavioral domain (i.e. negative or positive valence systems) yielded significant results (domain X direction of regulation; χ^2^ = 25.4, p=0.000042, domain X NF probe; χ^2^ = 33.4, p=0.000009, domain X instructions; χ^2^ = 34.2, p=0.000037). Together, the results suggest that the targeted neurobehavioral domain is the dominant consideration when designing Amygdala-NF experiments. In contrast grouping studies by the diagnostic category of the study’s population yielded no significant results (Figure 2c & Supplementary Figure 1, population X direction of regulation; χ^2^ = 7.3, p=0.5, population X NF probe; χ^2^ = 17.4, p=0.14, population X instructions; χ^2^ = 13, p=0.67), suggesting that the diagnostic category of the study’s population is not a prominent factor dictating design choices.

### 3. Neural mechanism of successful Amygdala self-modulation

Commonalities in BOLD activation across the different samples and designs (All participants’ activation; n=151) were analyzed by contrasting the *regulate* and *baseline* conditions (termed here *neuromodulation*) of a single NF session aimed to down modulate Amygdala activity (Figure 3a). This contrast revealed increased activation during *neuromodulation* in a distributed network including anterior insula (bilateral), lateral prefrontal cortex (bilateral), right lateral occipital cortex, supplementary motor area, and dorsal striatum (bilateral), as well as distributed decreased activation in the posterior insula and posterior cingulate cortex (PCC). (For the complete whole-brain activation results see supplementary Table 2).

**Figure 3.**
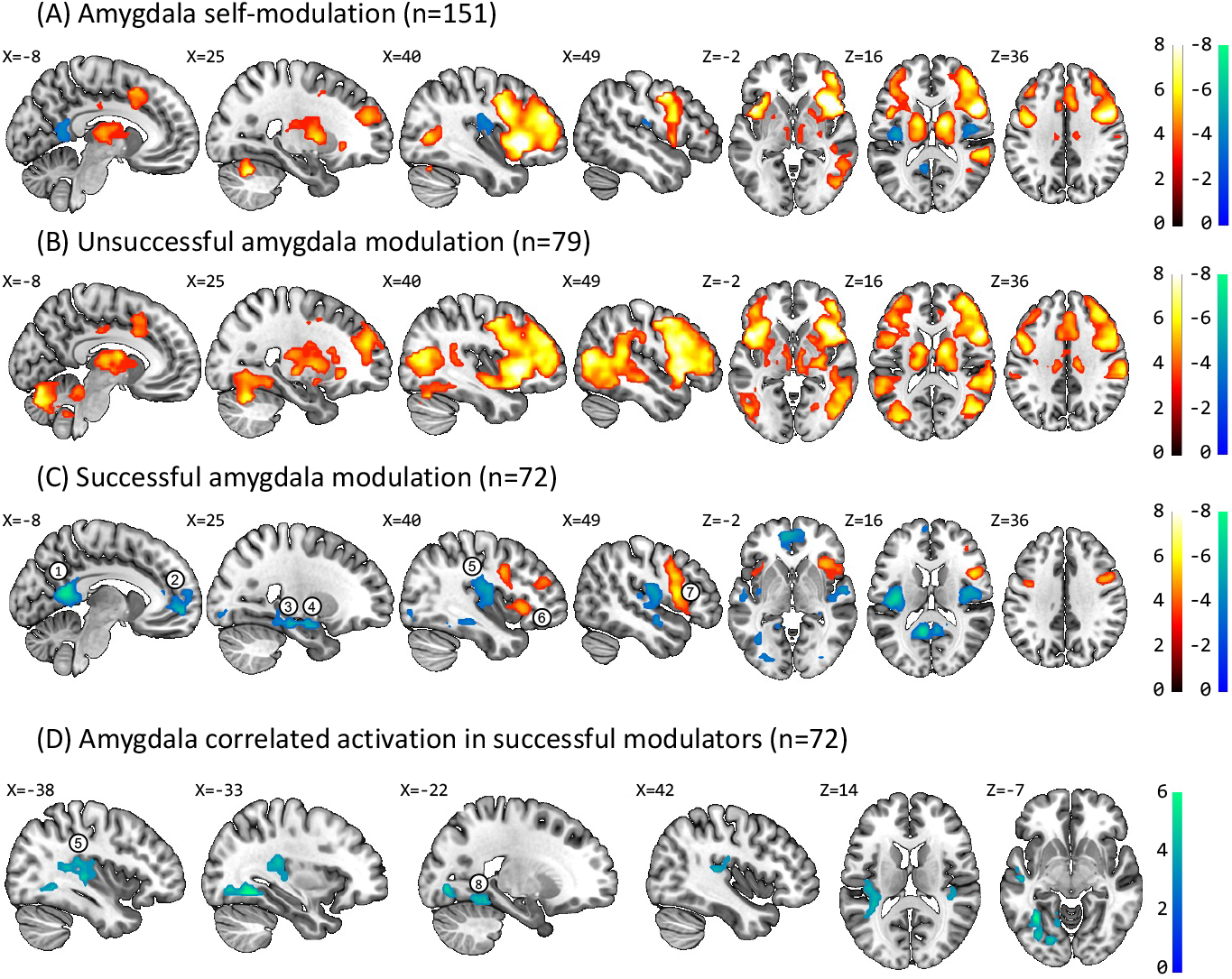
Whole brain analysis of Amygdala fMRI-NF for down-modulation. All images were assessed for cluster-wise significance at pfdr<0.05; cluster-defining threshold p=0.001; (A) All participants’ activation for regulate<baseline (n=151) showing activation pattern regardless of regulation success. Activations include positive clusters in the anterior insula (bilateral), lateral prefrontal cortex (bilateral), right lateral occipital cortex, supplementary motor area, and dorsal striatum (bilateral) and negative activation clusters in the posterior insulae and posterior cingulate cortex. (B) Activation pattern for Unsuccessful Amygdala modulation group (n=79, mean Amygdala beta=0.43, SD=0.4). Positive activations were observed in the anterior insula (bilateral), lateral prefrontal cortex (bilateral), lateral occipital cortex (bilateral), supplementary motor area, and dorsal Striatum (bilateral). (C) Activation pattern for Successful Amygdala modulation group (n=72, mean Amygdala beta=-0.51, SD=0.62). Decreased activation during regulate vs baseline was observed in the posterior cingulate cortex (marked 1), medial prefrontal cortex (marked 2), right hippocampus, and right Amygdala (marked 3,4) and dorsal posterior insula (marked 5). Increased activations during regulate vs baseline were observed in the anterior insula (marked 6), as well as in the lateral prefrontal cortex (marked 7). (D) Amygdala down modulation correlated activation (group level covariate) performed for successful modulators only (n=72). Down modulation of Amygdala activity positively correlated with posterior insula activity (marked 5) and left parahippocampal gyrus/fusiform gyrus (marked 8).

Analysis per “Successful” and “Unsuccessful” modulators revealed different activation patterns for the contrast of *regulate vs baseline* for each group of trainees. The group of “unsuccessful modulators” (n=79, defined as exhibiting an average Amygdala activity of greater than zero for *regulate vs baseline*; mean Amygdala beta=0.43, SD=0.4) showed distributed increased activations during *neuromodulation* in bilateral anterior insula, lateral PFC, lateral occipital cortex, supplementary motor area, and dorsal striatum (Figure 3b). The “successful modulators” group (n=79; defined as exhibiting an average Amygdala activity of less than zero for *regulate vs baseline;* mean Amygdala beta=-0.51, SD=0.62) showed a rather restricted activation during *neuromodulation* in bilateral anterior insula, lateral PFC, and wider spread negative activations most prominent in midline cortical regions (including ventro medial PFC and PCC), right hippocampus and bilateral posterior insula (Figure 3c). To further elucidate success related activations in a non-discrete manner we performed a post-hoc second level analysis with a calculated “success index” (average Amygdala activity for regulate vs baseline) used as a covariate. This analysis revealed co-variation in activity in bilateral posterior insula and left parahippocampal gyrus/fusiform gyrus (Figure 3d). Activation in these regions was correlated with neuromodulation success, so that these regions were negatively activated together with the Amygdala as a function of neuromodulation success.

## Discussion

Our triparted study provides a comprehensive overview and quantitative assessment of existing NF studies targeting Amygdala activity through three approaches: (a) a meta-analysis comprised of 20 studies examining effects of neuromodulation, learning and clinical outcome, (b) summary of design parameters and their association with valence system and (c) a multi-study fMRI mapping of successful down neuromodulation. The meta-analysis revealed that volitional Amygdala modulation is feasible and is a learnt skill (Figure 1, a-d), but its clinical utility is yet hard to evaluate due to parsed evidence from randomized placebo-controlled trials (Figure 1 e-f). The quantified distribution of design parameters across Amygdala NF studies demonstrated a large variety between studies on several parameters (Figure 2 a-b), though also pointed to a possible explainable framework of design choices around positive and negative valence systems (Figure 2c). fMRI analysis of a relatively large cohort suggested that successful down-modulators of Amygdala activity recruited a different brain network than unsuccessful modulators, consisting of mainly deactivation in the posterior insula and midline regions of the Default Mode Network (DMN; Figure 3c).

### Amygdala neuromodulation and learning effects

The meta-analysis of Amygdala-NF studies (including those using BOLD- and EFP probes) pointed to a significant *Neuromodulation effect* (*i.e. regulate vs baseline*) both within and between groups (Figure 1a-b). This suggests that Amygdala modulation cannot be attributed merely to general processes involved in NF training such as expectations or reward processing (Sorger et al., 2019). Yet, the sustainability of these modulation effects across time, beyond the training sessions, was not available for assessment and requires further consideration (Strehl, 2014). However, some studies using transfer testing with prospective fMRI (before and after training), showed greater ability to down regulate Amygdala activity in a different setting than the training (e.g. 15). Follow up testing also supported the notion that Amygdala-NF results in a sustainable effect that to a certain degree may be enhanced over time (Goldway et al., 2019; Rance et al., 2018). The large effect size pooled for the *Learning effect* (i.e. signal change in last vs first NF session) indicates that real feedback facilitates learning across time (Figure 1c-d). This finding supports the conceptualization of NF as a reinforcement learning process that could probably benefit from repeated training sessions (Sitaram et al., 2017). It is yet unclear, however, how many sessions are needed to achieve sufficient learning. In addition, it is reasonable to assume that different individuals would learn optimally trough different NF protocols (e.g. number/length of session) but additional investigation is needed. With respect to the clinical effect, our meta-analysis demonstrate that Amygdala-NF clinical research has not provided enough evidence to support a tangible conclusion regarding its clinical utility.

The present meta-analyses should be interpreted with caution since they suggest a substantial heterogeneity of effect sizes between studies. This is indicated by the values of parameter I^2^ (Higgins and Thompson, 2002) (see figure 1 a-f), illustrating that a large proportion of the total variation in the described sample remains to be explained. Interestingly, we tested whether the certain procedure parameter (i.e. population type, sample size, number of sessions or target valence system) might explain the observed differences in neuromodulation effect sizes. However, results of this analysis did not yield any statistical significance (see supplementary table 1), indicating that none of the above mentioned parameters could, by itself, explain the variability in modulation effect size. This suggests that the between-study heterogeneity may stem from more than one distinct variable, and perhaps, interaction between design choices.

### Characterization of variability in design parameters’ choice

An intriguing finding from the summary of design parameters was that the variation in design choices becomes more explainable when considering the targeted valence system (negative or positive), than when considering the study population (Figure 2c). In other words, researchers are in fact designing Amygdala-NF procedures in light of the RDOc valence system conceptualization, which is consisted within the rational of precision psychiatry. This finding supports the growing notion that treating abnormality in a specific *neurobehavioral process* might be beneficial cross-diagnostically, as such abnormalities are often impaired in multiple psychiatric diagnoses (Cuthbert, 2014). This conceptualization alluded to our recently proposed framework of process based NF (63). Under this framing, it seems reasonable that similar NF protocols are used to treat different psychopathologies as long as they target a common underlying process. For example, as revealed in Figure 2c, up-modulation of the Amygdala accompanied with positive valence affect is used to treat both MDD (24,25,39) and PTSD (Misaki et al., 2018b; Zotev et al., 2018) while NF protocols aimed at Amygdala down-modulation interfacing with negative valence, are used to treat PTSD (Fruchtman et al., 2019; Gerin et al., 2016; Nicholson et al., 2018), BDP (Paret et al., 2016a; Zaehringer et al., 2019) as well as chronic pain (Goldway et al., 2019). It might be first seen as paradoxical that “opposing” interventions such as up or down self-modulation of the Amygdala can be used to treat the same disorder. Taking as an example the case of PTSD, may help to clarify this point. Despite the fact that the hallmark symptoms of PTSD are manifested in the negative valance system (e.g. threat detection, fear learning and emotion regulation (Shalev et al., 2017)) it is also contributed by malfunctions in the positive valance system, such as reward related dysfunctions (e.g. anhedonia (Nawijn et al., 2015)). To this end, it should not be surprising that both down regulation of the Amygdala interfaced with negative context, and up-regulation Amygdala interfaced with a positive context, are effective in treating PTSD. To illustrate this point, Figure 4 exemplifies how two NF design aspects; feedback interface and outcome measures, when selected in a process-based manner, could potentially enhance precision in Amygdala-NF for PTSD. According to this proposal the impaired process characterization is guided by an assumed neuro-cognitive mechanism that underlies a certain symptom cluster in PTSD (e.g. avoidance vs hyperarousal) seemingly dominating the individual’s clinical phenotype. By applying such a framework it might be more feasible to establish an individually-tailored NF intervention for PTSD. Figure 4 demonstrates this point through three processes indicated in previous animal and human research as related to PTSD abnormalities (Fenster et al., 2018; Shalev et al., 2017): threat detection, emotion regulation, and fear extinction.

**Figure 4:**
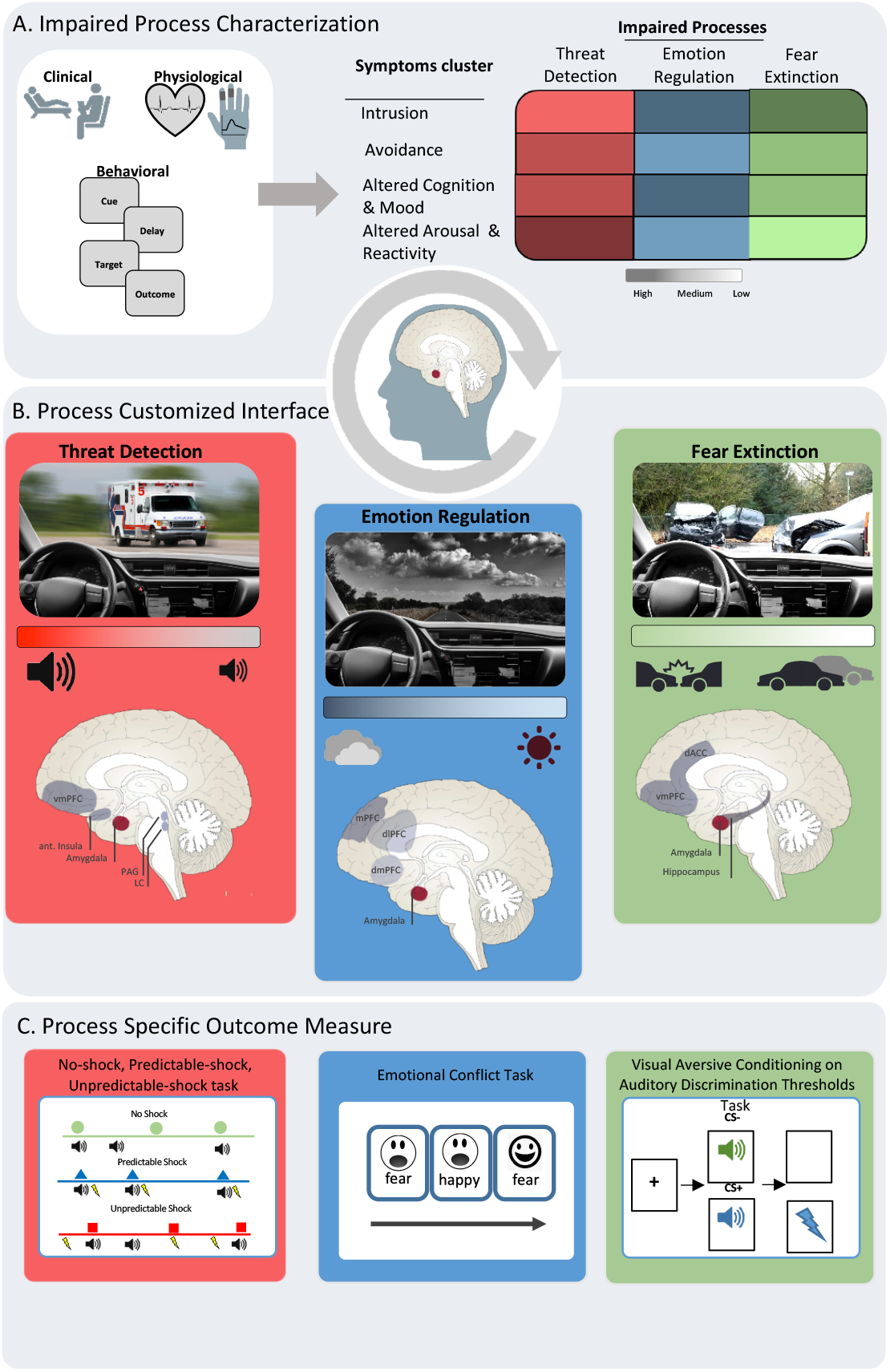
Suggested framework for Process-Based Amygdala-NF for PTSD (A). Impaired process characterization. Clinical intake, behavioral (i.e. questionnaires and process- specific tasks), and physiological (heart-rate and skin conductance during task) measures are used to characterize impaired process assessment in each patient. The impaired process characterization is guided by an assumed neuro-cognitive mechanism that underlies the main symptom clusters in PTSD. It is assumed that based on such assessment it is possible to establish an individually-tailored process-targeted NF intervention for PTSD. The table on the right demonstrates in a schematic fashion this idea with regard to three processes indicated in previous animal and human research as related to PTSD abnormalities (Fenster et al., 2018; Shalev et al., 2017): threat detection, emotion modulation, and fear extinction. PTSD symptom clusters (y-axis) are depicted in the table according to their suggested weights in each of the major dysfunctional processes (x-axis) per patient. (B). Individually-tailored process-based NF. The dysfunctional processes derived from the initial assessment battery will guide the selection of the corresponding intervention interface. Each interface is assumed to specifically target an impaired process by provoking activity in the designated brain circuitry alongside with the Amygdala (as shown by the brain illustrations) (suggested network are inspired by refs 83,84). It is further expected that Amygdala-NF in each unique context will yield specific modulation patterns of the underlying circuit of interest. For example: in the case of threat detection impairment, an interface with threat-related cues will be utilized, so that Amygdala activity feedback will correspond to the volume of an ambulance siren. Accordingly, this process-specific context will provoke modulation of threat detection related circuits involved in increased attention, reactivity to threatening stimuli, and hypervigilance, such as the anterior insula, vmPFC, periaqueductal gray, and locus coeruleus. (C). Process specific outcome measures. The change in the target process will be assessed using a designated behavioral paradigm. In this example, the “predictable and unpredictable shock task” (Abbott et al., 1984) for threat detection; “emotional conflict task” (Etkin et al., 2006) for emotion modulation and “visual aversive conditioning on auditory discrimination thresholds task” (Shalev et al., 2018) for fear extinction. This behavioral assessment will be done on top of clinical evaluation.

### Neural mediators of Amygdala self-modulation success

The cross labs’ fMRI analysis of Amygdala modulation (regulate vs baseline) during one NF session, revealed an expected pattern of activation including the anterior insula, lateral prefrontal cortex, supplementary motor area, and dorsal striatum corresponding to prior pooled analysis of NF studies (Emmert et al., 2016) (Figure 3a and supplementary Table 2). Further group analysis for successful and unsuccessful modulators revealed a distinct activation to each group (compare Figure 3b-c). The largely distributed activation in the unsuccessful modulators could imply that the NF-network activation suggested previously (Sitaram et al., 2017) may indicate an attempt to regulate neural signals and not necessarily successful neuromodulation (in this case; down regulating the Amygdala). A key finding from this grouped analysis is the deactivation (i.e. baseline *> regulate)* in a network that overlaps with the DMN (Raichle, 2015) in the group of successful modulators (Figure 3c). Of particular interest is the involvement of the vmPFC in successful neuromodulation as several Amygdala-NF studies have previously demonstrated changes in its functional connectivity after training (Keynan et al., 2019; Paret et al., 2016c; Zotev et al., 2013). The well-established connections of the vmPFC with the Amygdala (Ghashghaei et al., 2007), together with affect-regulation role of the DMN (Chiesa et al., 2013) suggests that multiple regulation processes contribute to Amygdala modulation such as; reappraisal (Urry et al., 2006), fear extinction (Phelps et al., 2004), and/or self-evaluation processes (Ochsner et al., 2005).

The second level covariance analysis within the “successful modulators” group, further showed deactivation (i.e. *baseline* > *regulate*) in the posterior insula that co-varied with Amygdala down-modulation. Presumably, this finding points to an involvement of introspection related processes in Amygdala-NF success. Indeed multiple neural pathways including the Amygdala and posterior insula were proposed to transmit information related to interception and somatosensorial, that were also linked to stress responses (McDONALD et al., 1999). Therefore, heightened anterior insula activation and reduced posterior insula activation among successful Amygdala modulators could reflect cognitive control over an interoceptive hub. Altogether the fMRI findings suggest that along with Amygdala down regulation there is a corresponding change in network activation that includes introspective and somatosensory processing.

To conclude, while our findings point to Amygdala self-modulation as a learned skill that could modify brain functionality that is trans-diagnostically related to mental illness, further placebo-controlled trials are necessary to prove clinical efficacy. We further suggest that studies should explicitly target neuro-behavioral processes, design the study accordingly and include ‘target engagement’ outcome measures rather than solely focusing on self-reported symptomatic change.

## Data Availability

All data produced in the present study are available upon reasonable request to the authors

## Acknowledgments

We are grateful for Tom Fruchtman-Steinbok for her help with the visual illustrations and comments. Prof. Hendler is a member of the BRAINTRAIN consortium, a collaborative project supported by the European Commission under the health cooperation work program of the 7th framework program, under grant agreement no. 602186. We would like to thank the following grants: US Department of Defense- grant agreement no. W81XWH-11-1- 0008. Mafat, IDF, I-Core cognitive studies grant agreement no. 693210. The Israeli Ministry of Science, Technology, and Space (Grant No. 3-11170). Kamin Program of the Israel Innovation Authority (Grant No. 59143), and the Sagol Network for Brain Research. The European Union’s Horizon 2020 research and innovation program supported this project under Grant Agreement No. 794395 to Lydia Hellrung. The funders had no role in study design, data collection, analysis, nor the publishing and preparation of this manuscript.

