## supplementary materials for "From Feasibility to Utility: A Meta-Analysis of Amygdala-Neurofeedback"

### Supplementary Material

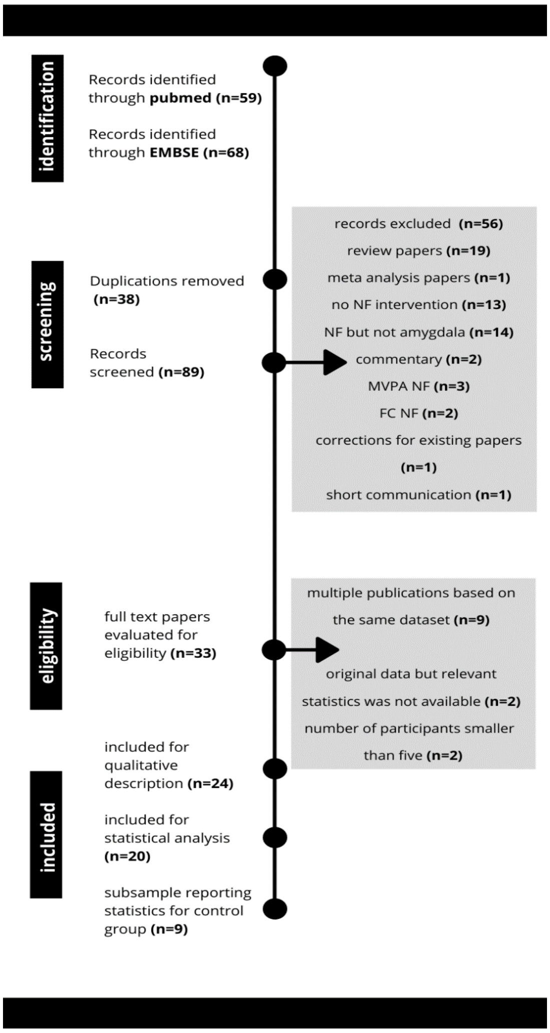


Supplementary Figure 1**:** Flow Diagram

**Subgroup analysis for "*neuromodulation*" effect size**

To evaluate whether procedural parameters (Target population, Instructions
Targeted system, Direction of regulation and Number of sessions) influenced NF effect size abilities, we performed a "subgroup analysis" (55) for the *Neuromodulation* effect as it included the largest number of studies. This analysis allowed to detect whether differences in study design may explain between-study variance of the estimated effect size. That is, subgroup analyses examine the studies of our meta-analysis to determine if they differ in terms of their effects.

To achieve maximal statistical power for this analysis we have clustered together subgroups across categories: For target population, we examine *healthy* vs *patients*; For instructions, we clustered *Positive Memories* with *Emotion regulation*; For target systems: positive vs negative valance, up vs down modulation and for number of sessions we used a slightly different method called meta regression (Borenstein et al., 2011). Results of this analysis, detailed in supplementary table 1, demonstrate that none of the design parameters had a significant contribution to the *neuromodulation* effect size.

| Factor | Q/F value | DF | p | | K | SDM | SE |
|---|---|---|---|---|---|---|---|
| Target population | 2.1 | 1 | 0.15 | | | | |
| | | | | Healthy | 12 | 0.68 | 0.1 |
| | | | | Patients | 8 | 1.14 | 0.31 |
| Instructions | 1.6 | 1 | 0.2 | | | | |
| | | | | Positive Memories | 5 | 0.76 | 0.059 |
| | | | | Emotion regulation | 13 | 1.02 | 0.19 |
| Targeted system | 0.16 | 1 | 0.68 | | | | |
| | | | | Negative valance | 13 | 0.87 | 0.14 |
| | | | | Pegative valance | 6 | 0.79 | 0.14 |
| Regulation Direction | 0.4 | 1 | 0.52 | | | | |
| | | | | Down | 12 | 0.91 | 0.14 |
| | | | | Up | 6 | 0.79 | 0.14 |
| # of sessions | 1.9 | 1,18 | 0.18 | | | | |

**Supplementary Table 1**: Parameters and their contribution to *neuromodulation* effect size. F values represent the statistic examining the difference between subgroups and the Q value, is being used in the case that continuous variable was used (i.e. number of sessions). K indicate the number of studies in each category SDM, summary measure for effect end SE is the standard error.

#### Methods for fMRI analysis

##### Included data

For the analysis of fMRI-NF, the authors focused on studies of amygdala downregulation in healthy individuals. The studies selected for analysis were studies where healthy subjects participated in a single session of fMRI-NF amygdala downregulation. We collected fMRI data from six studies, performed by three labs (detailed below). All subjects attempted to down-regulate their amygdalae (*regulate*), receiving visual feedback, and had a passive baseline condition in which similar visual stimuli were presented, without performing regulation nor receiving feedback (*view*). For datasets with control groups, only the experimental group data was used. For studies including both up- and down-regulation, only amygdala down-regulation data was used. Datasets differed in collection sites, scan parameters, and feedback protocols. All datasets were obtained as raw data (raw NIFTI or DICOM images) and were subjected to a uniform processing pipeline. Data consisted of 6 independent samples, 4 of which were previously published. All datasets are briefly described below, for more detailed descriptions, see original publications:

***Dataset 1*** (Paret et al., 2014)***:*** **collection site:** Central Institute of Mental Health, Mannheim, Germany **demographics:** n=16, Age=24.19, SD=4.17, 16 Females **target region:** Bilateral amygdala, 30% voxels with highest contrast estimate **study design:** Conditions: *regulate*, *view*, *neutral*; Interface: negative images (*regulate* & *view* conditions), scrambled images (*neutral*); Design: 3 runs * 5 cycles, trial length=24s **scan parameters:** Scanning was performed on a 3T Siemens Trio, TR=2000ms, TE=30ms, flip angle=80◦ 36 slices slice thickness: 3mm, slice gap: 1mm. **instruction:** Down-regulate (implicit)

***Dataset 2*** (Hellrung et al., 2018)***:*** **collection site:** Max Planck Institute for Human Cognitive and Brain Sciences, Leipzig, Germany **demographics:** n=34, Age=26.86, SD=3.14, 0 Females **target region:** Left amygdala **study design:** Conditions: Up-regulate, Down-regulate (*regulate*), *view*. Interface: Visual bar. One sub-group received continuous feedback (n=16), another group received intermittent feedback (n=18). Design: 3 practice runs * 4 blocks * 3 trials, trial length=40s **scan parameters:** Scanning was performed on a 3T Siemens MAGNETOM Trio. TR=2000ms, TE=25ms, flip angle=90◦, slice thickness=2.6mm, slice gap=0.26mm **instruction:** Down-regulation: count backwards from 100 subtracting 3, Up-regulation: positive memories.

***Dataset 3*** (Keynan et al., 2019) ***:*** **collection site:** Tel Aviv Medical Center, Israel **demographics:** n=56, Age=19.75, SD=1.35, 0 Females **target region:** Right amygdala **study**

**design:** Conditions: *regulate*, *view*. Interface: Flash animation of a boy skateboarding down a rural road. Feedback is given as the speed of the skateboard, which is displayed on a meter, and the participant's goal is to decrease the speed. One sub-group (n=30) practiced EEG EFP-NF before the fMRI session. Design: one subgroup competed 1 NF run * 2 cycles * 60s (n=17), a second subgroup competed 1 NF run * 2 cycles * 60s, and a second run of 5 cycles * 60s (n=39) **scan parameters:** Scanning was performed on a 3T Siemens PRISMA (n=44) and on a 3T GE Signa (n=12). On both scanners, NF runs were acquired with TR=3000ms, TE=35ms, flip angle=90◦, slice thickness=3mm, slice gap=0mm **instruction:** Down-regulate

***Dataset 4:*** **collection site:** Tel Aviv Medical Center, Israel **demographics:** n=9, Age=25.67, SD=4.37, 5 Females **target region:** Right amygdala **study design:** Conditions: *regulate*, *view*. Interface: Flash animation of a boy skateboarding down a rural road. Feedback is given as the speed of the skateboard, which is displayed on a meter, and the participant's goal is to decrease the speed.  1 NF run * 2 cycles * 60s, and a second run of 5 cycles * 60s scan **parameters:** Scanning was performed on a 3T Siemens PRISMA. TR=3000ms, TE=35ms, flip angle=90◦, slice thickness=3mm, slice gap=0mm **instruction:** Down-regulate. Previously unpublished data

***Dataset 5:*** **collection site:** Tel Aviv Medical Center, Israel **demographics:** n=27, Age=25.44, SD=3.12, 16 Females **target region:** Right amygdala **study design:** Conditions: *regulate*, *view*. Interface: Flash animation of a boy skateboarding down a rural road. Feedback is given as the speed of the skateboard, which is displayed on a meter, and the participant's goal is to decrease the speed. Design: 1 NF run * 3 cycles * 180s **scan parameters:** Scanning was performed on a Siemens PRISMA 3T. TR=3000ms, TE=35ms, flip angle=90◦, slice thickness=3mm, slice gap=0mm **instruction:** Down-regulate. Previously unpublished data

***Dataset 6*** (Keynan et al., 2016) ***:*** **collection site:** Tel Aviv Medical Center, Israel **demographics:** n=10, Age=25.3, SD=2.53, 5 Females **target region:** Right amygdala **study design:** Conditions: *regulate*, *view*. Interface: Flash animation of a boy skateboarding down a rural road. Feedback is given as the speed of the skateboard, which is displayed on a meter, and the participant's goal is to decrease the speed. Design: 1 NF run * 2 cycles * 60s, and a second run of 5 cycles * 60s **scan parameters:** Scanning was performed on a 3T GE Signa, TR=3000ms, TE=35ms, flip angle=90◦, slice thickness=3mm, slice gap=0mm **instruction:** Down-regulate.

**Supplementary Table 2**

**Amygdala modulation activity *Regulate*>*Baseline* (all participants n=151)**

| Cluster | Anatomical label | Volume (voxels) | p-FDR (cluster) | Activation direction | Hemisphere | x | y | z | peak p-unc. |
|---|---|---|---|---|---|---|---|---|---|
| 1 | Inferior Frontal Gyrus (pars oper) | 4505 | 0 | pos | R | 51 | 11 | 8 | 4.44E-16 |
| | Anterior Insula | | | pos | R | 33 | 20 | 5 | 4.44E-16 |
| | Middle Frontal Gyrus | | | pos | R | 51 | 8 | 29 | 4.44E-16 |
| | Anterior Insula | | | pos | L | -36 | 17 | -1 | 4.44E-15 |
| | Dorsal Striatum | | | pos | R | 15 | -4 | 17 | 2.52E-14 |
| | Precentral Gyrus | | | pos | L | -51 | -1 | 44 | 5.25E-14 |
| | Pallidum | | | pos | R | 18 | 5 | 2 | 1.74E-11 |
| | Caudate Nucleus | | | pos | L | -15 | -4 | 14 | 2.17E-11 |
| | Thalamus | | | pos | R | 6 | -22 | 5 | 1.32E-10 |
| 2 | Temporal Parietal Junction | 755 | 0 | pos | R | 63 | -37 | 20 | 5.42E-13 |
| | Lateral Occipital Cortex | | | pos | R | 48 | -67 | 8 | 2.63E-10 |
| 3 | Supplementary Motor Area | 428 | 0 | pos | midline | 3 | 14 | 47 | 7.89E-14 |
| 4 | Middle Cingulate Cortex | 64 | 0.014695 | pos | midline | 12 | -19 | 35 | 2.71E-06 |
| 5 | Cerebellum | 102 | 0.0032 | pos | R | 27 | -64 | -28 | 1.03E-13 |
| 6 | Parietal Operculum | 98 | 0.003245 | neg | R | 48 | -13 | 17 | 3.3E-06 |
| 7 | Posterior Insula | | | neg | R | 36 | -19 | 11 | 0.000116 |
| 8 | Posterior Cingulate Cortex | 67 | 0.014173 | neg | midline | -6 | -49 | 23 | 5.33E-06 |

**Unsuccessful amygdala modulation group Regulate > Baseline (n=79)**

| Cluster | Anatomical label | Volume (voxels) | p-FDR (cluster) | Activation direction | Hemisphere | x | y | z | peak p-unc. |
|---|---|---|---|---|---|---|---|---|---|
| 1 | Inferior Frontal Gyrus | 10880 | 0 | pos | R | 57 | 14 | 2 | 1.12E-14 |
| | Anterior Insula | | | pos | L | -45 | 11 | -1 | 1.74E-13 |
| | Cerebellum | | | pos | L | -30 | -67 | -25 | 4.2E-13 |
| | Anterior Insula | | | pos | R | 33 | 20 | 5 | 4.85E-13 |
| | Lateral Occipital Cortex | | | pos | R | 51 | -70 | 8 | 2.89E-12 |
| | Temporal Parietal junction | | | pos | R | 63 | -37 | 20 | 1.06E-11 |
| | Middle Frontal Gyrus | | | pos | R | 36 | 47 | 26 | 1.07E-11 |
| | Anterior Insula | | | pos | L | -36 | 17 | -1 | 1.43E-11 |
| | Precentral Gyrus | | | pos | R | 45 | -4 | 44 | 2.2E-11 |
| | Precentral Gyrus | | | pos | L | -45 | 2 | 41 | 3.34E-11 |
| | Thalamus | | | pos | R | 15 | -4 | 14 | 3.54E-11 |
| | Temporal Parietal Junction | | | pos | L | -51 | -43 | 23 | 1.19E-10 |
| | Cerebellum | | | pos | R | 30 | -61 | -28 | 1.55E-10 |
| | Thalamus | | | pos | L | -12 | -4 | 14 | 2.71E-10 |
| 2 | Supplementary Motor Area | 826 | 0 | pos | midline | 3 | 14 | 47 | 1.2E-11 |
| | Middle Cingulate Cortex | | | pos | midline | 0 | 11 | 29 | 6.8E-07 |

**Successful modulation group; Regulate>baseline (n=72)**

| Cluster | Anatomical label | Volume (voxels) | p-FDR (cluster) | Activation direction | Hemisphere | x | y | z | peak p-unc. |
|---|---|---|---|---|---|---|---|---|---|
| 1 | Inferior Frontal Gyrus | 488 | 0 | pos | R | 48 | 11 | 11 | 6.8E-09 |
| | Middle Frontal Gyrus | | | pos | R | 48 | 8 | 29 | 7.4E-08 |
| | Anterior Insula | | | pos | R | 33 | 20 | 2 | 8.61E-08 |
| 10 | Dorso-lateral Prefrontal Cortex | 50 | 0.020047 | pos | R | 39 | 41 | 23 | 7.81E-06 |
| 13 | Dorso-lateral Prefrontal Cortex | 35 | 0.04392 | pos | L | -42 | 29 | 26 | 2.17E-06 |
| 12 | Anterior Insula | 41 | 0.032202 | pos | L | -36 | 17 | -1 | 8.07E-06 |
| 2 | Superior Temporal Gyrus | 446 | 0 | neg | R | 54 | -4 | -13 | 6.66E-10 |
| 3 | Posterior Insula | 376 | 0 | neg | L | -45 | -22 | 17 | 3.3E-10 |
| 4 | Posterior Cingulate Cortex | 355 | 0 | neg | L | -9 | -55 | 11 | 1.57E-11 |
| 5 | Ventro-medial prefrontal cortex | 237 | 0.000003 | neg | midline | 0 | 56 | -7 | 5.63E-09 |
| 6 | Parahippocampal Gyrus | 229 | 0.000003 | neg | R | 27 | -25 | -19 | 2.72E-09 |
| 7 | Occipital Cortex | 166 | 0.000043 | neg | L | -21 | -94 | -10 | 2.04E-09 |
| 11 | Occipital Cortex | 46 | 0.024267 | neg | R | 33 | -85 | -13 | 5.62E-06 |
| 9 | Dorso-lateral Prefrontal Cortex | 69 | 0.006265 | neg | L | -21 | 26 | 44 | 1.12E-06 |
| 14 | Parahippocampal Gyrus | 35 | 0.04392 | neg | L | -24 | -37 | -13 | 5.15E-08 |

**Correlates of amygdala modulation; *Regulate>Baseline* with group covariate (n=72)**

| | | | | | | | | | |
|---|---|---|---|---|---|---|---|---|---|
| 1 | Superior Temporal Gyrus | 245 | 0.000007 | pos | L | -57 | -16 | 8 | 2.02E-06 |
| | Posterior Insula | | | pos | L | -36 | -28 | 5 | 2.87E-06 |
| 2 | Fusiform Gyrus | 196 | 0.00003 | pos | L | -30 | -55 | -13 | 1.05E-10 |
| | Parahippocampal Gyrus | | | pos | L | -24 | -46 | -16 | 3.89E-07 |
| 3 | Superior Temporal Gyrus | 57 | 0.038259 | pos | R | 57 | -19 | 8 | 1.5E-06 |

Borenstein, M., Hedges, L. V., Higgins, J. P. T., & Rothstein, H. R. (2011). *Introduction to Meta-Analysis*. John Wiley & Sons.

Hellrung, L., Dietrich, A., Hollmann, M., Pleger, B., Kalberlah, C., Roggenhofer, E., Villringer, A., & Horstmann, A. (2018). Intermittent compared to continuous real-time fMRI neurofeedback boosts control over amygdala activation. *NeuroImage*, *166*, 198–208.

Keynan, J. N., Cohen, A., Jackont, G., Green, N., Goldway, N., Davidov, A., Meir-Hasson, Y., Raz, G., Intrator, N., & Fruchter, E. (2019). Electrical fingerprint of the amygdala guides neurofeedback training for stress resilience. *Nature Human Behaviour*, *3*(1), 63.

Keynan, J. N., Meir-Hasson, Y., Gilam, G., Cohen, A., Jackont, G., Kinreich, S., Ikar, L., Or-Borichev, A., Etkin, A., Gyurak, A., Klovatch, I., Intrator, N., & Hendler, T. (2016). Limbic Activity Modulation Guided by Functional Magnetic Resonance Imaging–Inspired Electroencephalography Improves Implicit Emotion Regulation. *Biological Psychiatry*, *80*(6), 490–496. https://doi.org/10.1016/j.biopsych.2015.12.024

Paret, C., Kluetsch, R., Ruf, M., Demirakca, T., Hoesterey, S., Ende, G., & Schmahl, C. (2014). Down-regulation of amygdala activation with real-time fMRI neurofeedback in a healthy female sample. *Frontiers in Behavioral Neuroscience*, *8*. https://doi.org/10.3389/fnbeh.2014.00299
